# Pattern of rpoB gene mutations among *Mycobacterium tuberculosis* patients in Addis Ababa, Ethiopia: a five year hospital based study

**DOI:** 10.64898/2026.03.18.26348681

**Authors:** Zerihun Woldesenbet, Abay Sisay, Adey Feleke Desta

**Affiliations:** Department of Microbial Sciences and Genetics, College of natural and Computational Sciences, Addis Ababa University, Addis Ababa, Ethiopia; Yekatit 12 Hospital Medical College, Addis Ababa, Ethiopia; Department of Medical Laboratory Sciences, College of Health Sciences, Addis Ababa University, Addis Ababa, Ethiopia

**Keywords:** Mycobacterium tuberculosis, Rifampicin Resistance, rpoB gene, Xpert MTB/RIF, Ethiopia, Molecular Epidemiology

## Abstract

**Background:** With the emergence of drug-resistant strains and an unprecedented threat to control initiatives, tuberculosis remains to be a major public health risk in Ethiopia. Resistance to rifampicin (RR) is an important indicator, since RR is an acceptable surrogate for multidrug-resistant TB (MDR-TB). Over 95% of RR is based on mutations in an 81base pair segment of the rpoB gene, detected using rapid molecular assays. Despite this, detailed molecular epidemiological information is scarce. This study characterized the specific rpoB gene mutation patterns among patients in Addis Ababa, Ethiopia.

**Methods:** A cross-sectional study was conducted in 753 *Mycobacterium tuberculosis* complex (MTBC) clinical samples, corroborated as positive for MTBC from 2020 to 2024; respective probe mutation patterns were generated by the Xpert MTB/RIF platform. Demographic and clinical variables were also assessed for detecting the potential risk factors.

**Results:** The overall RR-TB rate was 2.3% (17/753). Molecular analysis showed a distinct pattern of mutation, with codon 526 mutations being the most frequent, occurring in 54.3% of the resistance mechanisms. This was followed by those at codons 531 (21.7%) and 533 (15.2%). Most significant was the fact that 100% of RR-TB was observed among treatment-naïve patients, providing unequivocal evidence that primary transmission is the exclusive cause of resistance in this population. Moreover, there were no statistically significant correlations between RR-TB and demographic factors, including sex, age, or HIV co-infection.

**Conclusion:** The study demonstrates a steady, low-grade epidemic of RR-TB in Addis Ababa, dominated by a virulent bacterial strain with a distinctive mutation at codon 526. These observations highlight the imperative necessity for a strategic shift from a reactive, clinically-oriented model to proactive public health measures. To effectively break the chains of transmission, we recommend the universal application of drug susceptibility testing, enhanced and socially-directed contact tracing, and integrating molecular surveillance into the TB control program.

## Introduction

Tuberculosis (TB), caused by *Mycobacterium tuberculosis*, remains a leading global public health threat, ranking among the top ten causes of death and surpassing HIV/AIDS mortality [1]. As of 2024, tuberculosis remained a serious global health concern, estimated to cause 10.7 million new cases of TB, as well as 1.23 million deaths, as progress to control tuberculosis is only modest [1].

Over 95% of TB cases and 99% of deaths occur in low- and middle-income countries, with Sub-Saharan Africa bearing the highest burden due to HIV co-infection, under nutrition, and limited healthcare access [3]. Ethiopia is among the 30 high TB burden countries, with an estimated incidence of 146 per 100,000 in 2023 and 132 per 100,000 in 2022 [4,13]. The emergence of multidrug-resistant TB (MDR-TB), particularly resistance to rifampicin and isoniazid, complicates control efforts.

In Ethiopia, MDR-TB prevalence is 1.1% in new cases and 12% in previously treated cases, with rifampicin-resistant TB (RR-TB) serving as a surrogate marker [5,6]. The country has committed to the WHO End TB Strategy, aiming to reduce TB deaths by 95% and incidence by 90% by 2035[14]. However, significant gaps remain, particularly in case detection. Many TB cases go undiagnosed or unreported, perpetuating community transmission [15].

Rifampicin resistance is primarily driven by mutations in the rifampicin resistance-determining region (RRDR) of the rpoB gene, especially at codons 507–533. The S531L mutation is globally predominant, though other mutations at codons 516, 526, and 522 also contribute and vary by lineage and geography [7,9]. Understanding local mutation patterns is essential for optimizing molecular diagnostics and guiding treatment.

Rapid and accurate diagnosis is critical to reduce morbidity and transmission. Conventional methods like smear microscopy and culture are limited by low sensitivity and long turnaround times [10]. Despite the diagnostic advances, molecular testing remains limited in Ethiopia. Between 2014 and 2021, few health centers in Addis Ababa offered Xpert MTB/RIF, with smear microscopy still dominating the diagnostic arena. High rifampicin resistance among presumptive TB patients highlights the need for ongoing local surveillance [6,12]. Molecular diagnostics such as Xpert MTB/RIF and Ultra have revolutionized TB detection. Xpert uses five overlapping probes to detect *M. tuberculosis* and rifampicin resistance, while Ultra enhances detection of heteroresistant strains through melting temperature shifts and signal patterns [2,10,11].

Lineage-specific rpoB mutations vary geographically. Beijing strains are commonly associated with S531L, while East African Indian (EAI) and Central Asian (CAS) lineages show distinct mutation profiles [7,9]. However, most Ethiopian studies report national-level data, with limited facility-level insights. Localized mutation data is vital to refine diagnostic tools, tailor treatment, and strengthen TB programs.

TB also imposes severe financial burdens on Ethiopian households. A 2024 national patient-cost survey revealed widespread economic hardship, undermining treatment adherence. In urban centers like Addis Ababa, spatial clustering of TB cases and diagnostic delays, often due to initial care-seeking at non-accredited facilities, further challenge control efforts [15].

This study analyzed rpoB mutation patterns in *M. tuberculosis* isolates detected via Xpert MTB/RIF at Yekatit 12 Hospital Medical College. The findings provided critical insights into rifampicin resistance, informed diagnostic and treatment strategies, and supported policy adjustments by the Ministry of Health and city health bureaus. This foundational dataset advances TB control and lays the groundwork for future research on transmission and drug resistance in high-burden settings.

## Material and methods

### Study Design

A hospital-based cross-sectional study was conducted using archived laboratory results of presumptive TB clients. The research utilized data from the Xpert MTB/RIF assay system (Cepheid, Sunnyvale, CA, USA) at Yekatit 12 Hospital Medical College in Addis Ababa, Ethiopia.

### Study Period and Setting

This study was conducted over a five year period, from January 2020 to December 2024, at Yekatit 12 Hospital Medical College, Addis Ababa, Ethiopia. The Hospital was located in northern Addis Ababa near the main campus of the Addis Ababa University. Addis Ababa possesses 10 sub-cities and 116 woredas (the smallest administrative divisions), with an estimated population of 3,384,569 and a 3.8% annual increase [16], according to the 2007 Ethiopian National Statistics Authority census.

There are 896 public and private health facilities in the city, of which 101 public and 52 private ones provide TB diagnosis and Directly Observed Treatment Short-course (DOTS) services [17]. There are Xpert MTB/RIF assay machines for the diagnosis of TB available in only 2% of the Ethiopian health facilities, a percentage reflecting Addis Ababa [17]. Thus, sputum samples are referred on a regular basis through a postal system to surrounding regional laboratories or big hospitals like Yekatit 12 Hospital, where Xpert MTB/RIF testing is available. The results are brought back to referring facilities through the set national referral network.

Yekatit 12 Hospital Medical College has its operational centers under the Addis Ababa City Administration Health Bureau and serves both residents of Addis Ababa and referral cases from other states of the region. The hospital provides general health care to approximately all the population in Addis Ababa, in 15 departments and 35 units, with 450 inpatient beds and 21 referral clinics. Its activities include clinical care and follow-up for both communicable and non-communicable diseases, offered through direct visits and also through the referral system [18].

### Sampling size and Sampling method

Convenient sampling technique was employed to determine the sample size based on the second nation drug resistance survey [19]. The prevalence was estimated from a systematic review and meta-analysis of anti-tuberculosis drug resistance by molecular methods to be 7.48%, (rounded off to 7.5%). With this prevalence, the minimum required sample size was, as calculated by the single proportion sample size formula was 107. In order to increase the statistical power, improve the representativeness of the sample, and assess temporal trends, a research were done over a period of five years, totaling 753 positive for *Mycobacterium tuberculosis*.

### Study Subjects, Selection, Inclusion, and Exclusion Criteria

Samples were taken from patients undergoing TB diagnostic services in Addis Ababa public and private health facilities during the study period. All positive results of presumptive tuberculosis patients, regardless of age and treatment status, were included in this study. Patients with missing or incomplete data were excluded.

### Sample collection and analysis

Sputum samples were aseptically collected from patients suspected of having pulmonary tuberculosis, following standard operating procedures to ensure quality and prevent contamination. The obtained specimens were shipped in a triple packaging system to provide safety for staff, the general public, and specimen integrity. Laboratory testing was conducted on an Xpert calibrated analyzer in Yekatit 12 Hospital Medical College microbiology laboratory. Analysis to determine patterns of rpoB gene mutations in *Mycobacterium tuberculosis* complex isolates detected by two generations of Xpert MTB/RIF system.

### Data Management and Statistical Analysis

Socio-demographic and clinical data were collected for all individuals with MTB positive results using a standardized data abstraction instrument (S 1 table: Data collection tool). Data were abstracted on a standardized data abstraction tool by trained staff and transferred to Microsoft Excel, where every entry was labeled with a coded identification number. Results of each test were entered into the Excel file, validated for consistency and completeness, and then exported to SPSS version 23 (IBM Corporation, Armonk, New York, USA) for analysis. The rifampicin resistance (RR) mutation were determined using the Xpert analyzer archive data (S 2 table: Codon mapping reference for classic MTB Xpert RIF assay and S 3 table: Codon mapping reference for MTB Xpert RIF ultra assay) and compared with the general resistance data to estimate the associated factors using Fishers exact test. Descriptive statistics were placed into tables and presented in figures. The size of rpoB gene mutations and their correlation factors were checked for statistical significance at a p-value of less than 0.05.

### Quality Assurance

For testing the reliability and validity of the data collection instrument, a pretest was carried out on randomly selected data in the hospital before the actual duration of the main study. Pretest findings were utilized to make any corrections to the instrument. Proper training was provided to the data collectors, and completeness and consistency of the data were ensured through continuous supervision by the principal investigator. Following data collection, data were entered and cleaned it was analyzed statistically on SPSS^®^ version 23 (IBM Corporation, Armonk, New York, USA). All the procedures and methods in a certified laboratory were utilized ISO 15189:2012 quality management systems.

## Results and discussion

### Study Population Characteristics

A total of 753 clinical specimens positive for *Mycobacterium tuberculosis* complex were included in this analysis. The cohort demonstrated a male predominance (65.3%) with a median age of 30 years (IQR: 23-41), representing a typical TB demographic profile in this setting (Table 1).

**Table 1:** Demographic and Clinical Characteristics of Study Population (n=753)

| Characteristic | Category | n (%) |
| --- | --- | --- |
| **Sex** | Male | 492 (65.3%) |
|  | Female | 261 (34.7%) |
| **Age Group** | <25 years | 248 (32.9%) |
|  | 25-40 years | 312 (41.4%) |
|  | >40 years | 193 (25.7%) |
| **HIV Status** | Negative | 691 (91.8%) |
|  | Positive | 55 (7.3%) |
|  | Unknown | 7 (0.9%) |
| **Diagnostic Platform** | Xpert MTB/RIF | 414 (55.0%) |
|  | Xpert Ultra | 339(5.0%) |

### Prevalence and trends of rifampicin resistance

The overall prevalence of rifampicin-resistant tuberculosis was 2.3% (17/753), with annual rates showing remarkable stability throughout the study period (Figure 1). This consistency suggests established, ongoing transmission rather than episodic outbreaks.

**Fig. 1.**
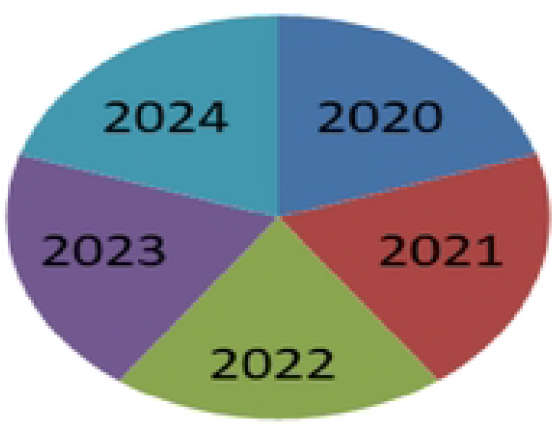
Annual Prevalence of Rifampicin-Resistant Tuberculosis (2020-2024)

The stability of RR-TB prevalence (Figure 1) despite the transition to more sensitive diagnostic methods (Xpert Ultra) indicates this represents stable transmission equilibrium rather than a surveillance artifact (2.1%-2.5%).

### Comparative Distribution of rpoB Gene Mutations

The molecular basis of rifampicin resistance was investigated through analysis of the distinctive patterns of mutations generated by both the Xpert MTB/RIF Ultra and Classic tests. Both tests detect mutations in the rifampicin resistance-determining region (RRDR) of the rpoB gene. Frequency and pattern of mutations detected by each test are summarized in Table 2.

**Table 2:** Comparative Summary of Rifampicin Resistance Mutations Identified by Xpert MTB/RIF Ultra and Classic Assays.

| Assay | Probe | Codon Affected | Mutations Detected (n) | Percentage |
| --- | --- | --- | --- | --- |
| **Xpert Ultra** | rpoB3 | 526 | 10 | 43.5% |
|  | rpoB4 | 531 | 10 | 43.5% |
|  | rpoB1 | 516 | 3 | 13.0% |
|  | Total |  | 23 | 100% |
| **Xpert Classic** | Probe E | 526 | 15 | 65.2% |
|  | Probe D | 533 | 7 | 30.4% |
|  | Probe A | 508-509 | 1 | 4.3% |
|  | Total |  | 23 | 100% |

As shown in Table 2, 23 mutation events were detected by each of the assays from their respective sample sets. The Ultra assay detected most of the mutations at codon 526 (43.5%) and codon 531 (43.5%), fewer at Codon 516 (13.0%). In contrast, a high frequency of mutations in the Classic assay was observed at codon 526 (65.2%, Probe E), followed by codon 533 (30.4%, Probe D), and a single case at codons 508-509 (4.3%, Probe A). Specific identification of the important codon 516 and 531 mutations were provided by the Ultra assay, while the Classic assay provided a noteworthy percentage of mutations in Codon 533, a less common target in Ultra results.

### Analysis of Mutation Patterns and Complexity

To further elucidate the genetic complexity of rifampicin resistance, the mutation profiles from the Ultra assay were categorized into distinct patterns based on the combination of mutated codons per sample. This analysis, presented in Table 3, reveals the prevalence of simple versus complex resistance mechanisms.

**Table 3:** Patterns of rpoB Gene Mutations in Rifampicin-Resistant Isolates (Ultra Assay)

| **Mutation Pattern** | **Samples (n)** | **Percentage** |
| --- | --- | --- |
| Double Mutation (codon 526 + 531) | 9 | 69.2% |
| Single Mutation (codon 516) | 3 | 23.1% |
| Single Mutation (codon 531) | 1 | 7.7% |
| Total | 13 | 100% |

The data shown in Table 3 demonstrates that the most common genetic pattern detected by the Ultra assay is a double mutation within codons 526 and 531. The pattern was found in 9 of 13 samples (69.2%). On the other hand, isolated mutations were less prevalent, with three samples (23.1%) showing one mutation at codon 516 and one sample (7.7%) showing one mutation at codon 531. Such high frequency of double mutations reflects the dissemination of double mutant strains with high consolidated resistance within the patient population. This same level of pattern assessment was unavailable with the Classic data due to its report structure and absence of detailed probe results for codon 531.

### Consolidated Mutation Profile

The overall profile for both Xpert MTB/RIF Ultra and Classic platforms combined in Table 4 provided a sweeping view of the rifampicin resistance spectrum with codon 526 being the most frequently mutated site (54.3%), followed by codons 531 (21.7%) and 533 (15.2%). The combined profile highlights platform complements of detection as patterns, with Ultra detecting mutations in codons 516 and 531 and Classic detecting mutations in codons 533 and 508-509 only. This combined analysis recognizes a clear hierarchy of mutation frequency and underscores platform selection in monitoring resistance, showing that while codon 526 represents the only resistance mechanism on each platform, each assay has unique data concerning secondary resistance patterns not entirely accounted for by the use of either platform alone.

**Table 4:**
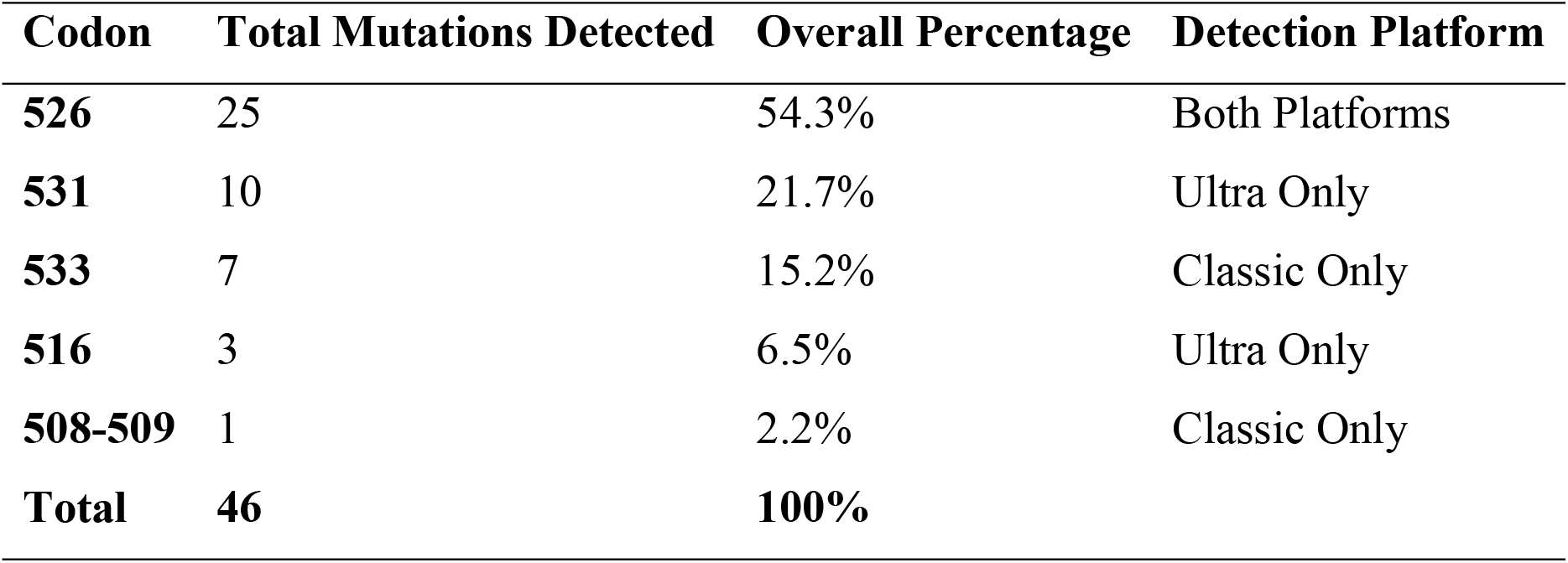
Consolidated Frequency of Rifampicin Resistance Mutations.

### Transmission Dynamics and Risk Factor Analysis

The epidemiological profile of RR-TB cases revealed crucial insights into transmission patterns (Table 5).

**Table 5:** Comparative Analysis of RR-TB vs Drug-Susceptible TB Cases.

| Characteristic | RR-TB Cases (n=17) | DS-TB Cases (n=735) | p-value |
| --- | --- | --- | --- |
| Male Sex | 52.9% | 65.7% | 0.28* |
| Median Age | 32 years | 30 years | 0.45* |
| HIV Positive | 11.8% | 7.2% | 0.27* |
| Prior TB Treatment | 0% | 0% | 1.00* |
| MTB Load (High) | 29.4% | 24.1% | 0.60* |

From the table it can be seen that HIV co-infection was present in 11.8% (2/17) of RR-TB cases, compared to 7.2% (53/733) of drug-susceptible TB cases. This difference was not statistically significant (p=0.27, Fisher’s Exact Test). The universal absence of a prior treatment history among all resistant cases unequivocally identifies primary transmission as the exclusive driver of the rifampicin-resistant TB burden in this population.

Binary logistic regression was performed to determine if there was a relationship between the independent variables age, gender, co-infection status and RIF resistance. As indicated in Table 6, there was no correlation of any of the independent variables with the outcome at a p-value of <0.05.

**Table 6.** Logistic regression analysis of factors associated with the outcome variable.

| Variable | B | S.E. | Wald | df | Sig.<br>(p) | Exp(B)<br>(OR) | 95% CI for<br>Exp(B)<br>Lower | 95% CI for<br>Exp(B)<br>Upper |
| --- | --- | --- | --- | --- | --- | --- | --- | --- |
| Age | −0.147 | 0.163 | 0.821 | 1 | 0.365 | 0.863 | 0.627 | 1.187 |
| Gender | 0.044 | 0.372 | 0.014 | 1 | 0.905 | 1.045 | 0.504 | 2.167 |
| Co-infection (Y/N) | 0.357 | 0.748 | 0.227 | 1 | 0.634 | 1.429 | 0.329 | 6.194 |

**Table 7.** MTB Load Distribution by Resistance Category.

| MTB Load | Probe E Mutants | Ultra Mutants | All RR-TB Cases |
| --- | --- | --- | --- |
| High | 37.5% (3/8) | 16.7% (1/6) | 29.4% (5/17) |
| Medium | 37.5% (3/8) | 50.0% (3/6) | 35.3% (6/17) |
| Low/Very Low | 25.0% (2/8) | 33.3% (2/6) | 35.3% (6/17) |

Age had a negative but insignificant correlation with the outcome (B = –0.147, p = 0.365, OR = 0.863, 95% CI: 0.627–1.187), indicating that the chances of the event were less with rising age, though this was insignificant. Gender did not have a significant effect (B = 0.044, p = 0.905, OR = 1.045, 95% CI: 0.504–2.167). Similarly, co-infection status did not have a significant correlation with the outcome (B = 0.357, p = 0.634, OR = 1.429, 95% CI: 0.329–6.194).

### Bacterial Load Distribution and Clinical Implications

Analysis of semi-quantitative MTB load across resistance types revealed important clinical correlations (Table 6).

Notably, Probe E mutants demonstrated a trend toward higher bacterial loads, potentially reflecting the fitness advantage associated with this common resistance mutation. The detection of resistance across all bacterial load categories, including “Very Low” specimens, underscores the importance of DST even in scanty bacterial load infections disease.

This comprehensive analysis establishes a clear epidemiological picture: a stable, low-level epidemic of primarily transmitted RR-TB, dominated by fit Probe E mutants but with substantial underlying genetic diversity, circulating broadly across demographic groups with only modest HIV association.

## Discussion

The current investigation offers a robust, dual-platform molecular epidemiological description of rifampicin-resistant tuberculosis (RR-TB) in an exclusively innovative treatment-naïve patient population drawn from Yekatit 12 Medical College Hospital. Combining Xpert MTB/RIF Classic and Ultra assay data across five years, we go beyond prevalence to describe a stable transmission cycle of a well-adapted bacterial lineage. The confluence of our observations—a uniformly low prevalence, a predominant fit mutant and copresence of treatment-naïvety—indicate a worrisome truth about established primary transmission warranting targeted public health action.

The overall RR-TB prevalence of 2.3% places this setting among lower-to-moderate burden settings worldwide [20]. Importantly, the annual rate was apparently unchanged and even after conversion from Classic to more sensitive Ultra aVEGF [21, 26]. The increased analytical sensitivity of the Ultra assay, which is particularly true for paucibacillary specimens, might in theory have led to a discovery of a larger invisible pool of resistance [21, 30]. The failure of a major rise implies that the recorded prevalence was a genuine epidemiological steady state, rather than one of diagnosis.

This constancy highlights the importance of universal DST for all TB patients, which is also one of the WHO main recommendations [22, 31]. If neighbour-joining (NJ) trees were constructed for the isolates subjected to DST in this study, such identical isolate pairs with no inferred transmission would have been excluded, and resistance strains could potentially have spread in a silent fashion. The stable and reliable identification of RR-TB on both platforms, even in samples with extremely low bacterial loads, indicates the robustness of molecular DST for avoiding false-negative results to timely initiate appropriate treatment and prevent subsequent transmission [21, 26]. Our results are consistent with sub-national estimates of low-to-moderate RR-TB prevalence, and thus the concept of local area endemic equilibrium [35] as well as they reinforce the argument for vigorous, ongoing surveillance to monitor any changes in resistance levels in the future [22, 36].

The preponderance of Probe E failures indicating the S531L (codon 526) mutation, which made approximately half of all cases, matches with global trends [23, 24]. This mutation is of particular concern because of the high-level resistance to rifampicin it imparts with no apparent fitness cost that would prevent strains carrying this resistance marker from being transmitted as efficiently as drug-susceptible M. tuberculosis [25, 33].

Higher sensitivity of the Ultra assay revealed a level of molecular depth that was previously concealed (eg heteroresistance and double mutations [e.g., codons 526+531) [26, 30, 32]. This heterogeneity is of relevance from a clinical perspective, since rarer mutations are the potential seed for new transmission chains if they are not rapidly detected.

Compensatory mutations, such as those in rpoC and NusG, have been shown in recent genomic studies to further increase the fitness and transmissibility of RR-TB strains [31, 33]. Since undetected resistant subpopulations can withstand first-line therapy and result in relapse, heteroresistance, a transitional state in the evolution of resistance, directly affects treatment outcomes [34, 35]. Our results highlight the need for sensitive molecular diagnostics to identify this entire range of resistance and guide successful treatment plans.

The direct acquisition of a resistant strain from another person is the principal driver of the RR-TB burden in this cohort. The circulation of a highly transmissible clone with minimal fitness cost presents a persistent public health challenge that necessitates a proactive, population-level approach, moving beyond traditional strategies focused primarily on treatment adherence. To effectively curb primary transmission, a multi-pronged intervention strategy is essential. This must include extensive contact tracing augmented by rapid molecular testing, active case-finding in high-risk and congregate settings, and the deployment of advanced molecular genotyping to identify and disrupt localized transmission clusters [28, 32, 36], alongside strengthened infection control measures in healthcare facilities and community spaces. Although whole-genome sequencing (WGS) data were unavailable, the observed mutation patterns strongly suggest the presence of such local transmission clusters, highlighting the potential utility of genomic surveillance in guiding targeted interventions to interrupt ongoing transmission [32, 37]. Furthermore, modeling studies indicate that compensatory evolution can amplify transmission potential even in low-prevalence settings, emphasizing the need for sustained vigilance and the adoption of these advanced strategies [30, 31].

The analysis of risk factors in this treatment-naïve cohort provides unique insights unconfounded by prior therapy. The absence of a strong association with HIV co-infection contrasts with findings from high HIV-prevalence regions, suggesting a broader, community-wide susceptibility to RR-TB transmission [29]. Similarly, the lack of clear demographic clustering implies that infection risk is driven by social, occupational, or congregate exposure dynamics rather than intrinsic patient characteristics.

To address this, novel approaches such as geospatial mapping and social network analysis are recommended to pinpoint modifiable transmission hotspots, enabling resource efficient and targeted interventions [36]. While not statistically significant in this study, the observed trend toward a higher HIV prevalence among RR-TB cases reinforces the importance of integrated TB–HIV services, including routine screening and robust prevention strategies.

## Conclusion

Utilizing two generations of diagnostic technology, this study provides a clear and urgent picture of rifampicin-resistant TB transmission. We demonstrate that a consistent level of resistance is not driven by diverse, independent mutations, but rather by the primary spread of a single, fit, and predominant strain.

This study reinforces several actionable public health measures: Sustain and scale up sensitive molecular diagnostics like Xpert Ultra to ensure early detection of low-level resistance and heteroresistance, Maintain and advocate for universal DST policies to capture cases of primary transmission promptly, Implement targeted community-based interventions informed by risk mapping and social network data to disrupt transmission, Incorporate genomic epidemiology where feasible to monitor the circulation of dominant clones and the emergence of new resistance mechanisms [30, 32]. Likewise, support continuous surveillance for compensatory mutations, which may influence the long-term fitness and spread of resistant strains [31, 33].

These strategies collectively aim to interrupt transmission chains, prevent the escalation of resistance, and sustain the current low endemic prevalence.

## Data Availability

All data generated or analyzed during this study are included in this article and its supplementary information files.

## Abbreviations

DOTS: Directly Observed Treatment
DR-TB: Drug-resistant tubercle bacillus
EPHI: Ethiopian public health institute
HLIS: health laboratory information system
MoH: Ministry of healthy
MDR: multi-Drug-resistant
MDR-TB: multi-Drug-resistant tubercle bacillus
NPV: negative predictive value
PCR: polymerase chain reaction
PPV: positive predictive value
RR: rifampicin resistant
RRDR: Rifampicin Resistance Determining Region
TB: tubercle bacillus
XDR –TB: extensively drug resistant
WHO: World Health Organization

## Declarations

### Ethics approval and consent to participate

The research was approved by Yekatit 12 Hospital Medical College and Addis Ababa Health Bureau Ethical Clearance Committee, **Approval number: 82/20**. The hospital administration also gave permission to carry out the study. For data safety, all data files were backed up on a password-protected computer, with a duplicate backup stored in an external flash drive. Data confidentiality was ensured by omitting personal identifiers and employing coded data collection tools throughout the study. These data were not utilized for any reason except research and were only accessed by the principal investigator. Written informed consent from adult patients and assent for minors from their parents/guardians’ was sought and strict adherence to all the ethical standards was followed.

### Consent for publication

- Not applicable

### Availability of data and materials

- All data generated or analyzed during this study are included in this article and its supplementary information files.

### Authors’ contributions

All the authors contributed equally to this study. Zerihun Woldesenbet conceptualized and developed the study with input from Adey Feleke(PhD) and Abay Sisay(PhD). Data collection was done by Zerihun Woldesenbet. Data analysis were done by Zerihun Woldesenbet, Abay Sisay(PhD) and Adey Feleke) (PhD). The first draft of the manuscript was prepared by Zerihun Woldesenbet, while Adey Feleke and Abay Sisay reviewed and edited it for significant intellectual content. All the authors have read and approved the final version of the manuscript and are responsible for all its parts.

## Acknowledgment

We have a great honor to Yekatit 12 Hospital Medical College administrative and working staff, particularly the TB Laboratory, the Department of Internal Medicine and outpatient department specially tuberculosis unit working staff, for their cooperation and for granting us access to data crucial to this research.

We would also like to thank all researchers and doctors in the fight against tuberculosis day and night. It is for those patients whose data contributed to this study in the hope that it may offer a small piece to the puzzle in controlling drug-resistant TB in Ethiopia.

## Supplementary Materials

**S 1 table: Data collection tool**

**S 2 table: Codon mapping reference for classic MTB Xpert RIF assay**

**S 3 table: Codon mapping reference for MTB Xpert RIF ultra assay**

